# Higher risk of SARS-CoV-2 Omicron BA.4/5 infection than of BA.2 infection after previous BA.1 infection, the Netherlands, 2 May to 24 July 2022

**DOI:** 10.1101/2022.09.21.22280189

**Authors:** Stijn P. Andeweg, Brechje de Gier, Harry Vennema, Ivo van Walle, Noortje van Maarseveen, Nina E. Kusters, Hester E. de Melker, Susan J.M. Hahné, Susan van den Hof, Dirk Eggink, Mirjam J. Knol

**Affiliations:** Center for Infectious Disease Control, WHO COVID-19 reference laboratory, National Institute for Public Health and the Environment (RIVM), Bilthoven, The Netherlands; Department of Medical Microbiology, University Medical Center Utrecht, Utrecht, the Netherlands; Saltro Diagnostic Center for Primary Care, Utrecht, The Netherlands

**Author notes:** Corresponding authors: Dirk Eggink and Mirjam J. Knol. These authors contributed equally to this article and share last authorship (Dirk Eggink, Mirjam J. Knol).

## Abstract

We investigate differences in protection from previous infection and/or vaccination against infection with Omicron BA.4/5 or BA.2. We observed a higher percentage of registered previous SARS-CoV-2 infections among 19836 persons infected with Omicron BA.4/5 compared to 7052 persons infected with BA.2 (31.3% vs. 20.0%) between 2 May and 24 July 2022 (adjusted odds ratio (aOR) for testing week, age group and sex: 1.4 (95%CI: 1.3-1.5)). No difference was observed in the distribution of vaccination status between BA.2 and BA.4/5 cases (aOR: 1.1 for primary and booster vaccination). Among reinfections, those newly infected with BA4/5 had a shorter interval between infections and the previous infection was more often caused by BA.1, compared to those newly infected with BA.2 (aOR: 1.9 (1.5-2.6). This suggests immunity induced by BA.1 is less effective against a BA.4/5 infection than against a BA.2 infection.

## Background

The severe acute respiratory syndrome coronavirus 2 (SARS-CoV-2) Omicron variants have led to large numbers of infections globally[1], driven by increased transmissibility and escape from vaccine- and infection-induced immunity[2-4]. Sub-variants of Omicron, mainly BA.1, BA.2, and BA.5, have been circulating globally. In the Netherlands, as in the rest of Europe, an initial BA.1 surge was observed starting at the end of 2021, with rapid succession by BA.2 in early 2022, followed by BA.4 and BA.5 mid 2022[5]. Omicron variants have a large number of substitutions in the spike protein compared to earlier variants of concern (VOC), and also between different Omicron lineages substantial differences in the spike protein are present[6]. All Omicron variants show reduced sensitivity to antibodies induced by vaccination, previous infection or both (hybrid immunity). The substitutions in BA.4/5 show the largest reduction of neutralization[7-9], raising concern about the protection by vaccination and/or previous infection, including protection conferred by previous infections by other Omicron lineages.

To examine a possible reduction in protection from vaccine- and/or infection-induced immunity against BA.4/5 infection compared to protection against BA.2 infection, we employ a case-only approach in which we study the effect of pre-infection immune status (based on previous infection and/or vaccination) on the occurrence of BA.4/5 and BA.2 during the transition period from BA.2 to BA.4/5 (2 May to 24 July 2022). In addition, we study reinfections to assess (1) whether the interval between current and previous SARS-CoV-2 infection differs by variant and (2) to assess differences in variants of the previous infections in newly infected BA.4/5 and BA.2 cases.

## Methods

### Epidemiologic data

From 1 June 2020 onwards, mass community testing for SARS-CoV-2 organized by the 25 regional Public Health Services has been available and advised for Dutch citizens experiencing COVID-19 like symptoms. Since 11 April 2022, mostly individuals at high-risk of severe disease and healthcare workers are advised to still visit the PHS for PCR testing. In the current study, we used positive tests from national SARS-CoV-2 community testing from 2 May to 24 July 2022 which were analyzed with the TaqPath COVID-19 RT-PCR kit (ThermoFisher Scientific, Nieuwegein, The Netherlands) to assess S-gene target failure (SGTF, see below). Of persons having multiple positive SGTF tests within 30 days during the study period, only the first positive test was included. Otherwise, both tests were included (this was only applicable to one individual).

Test results were linked to the national community testing register (CoronIT) using a unique sample number. The national community testing register contains pseudonymized data with demographic characteristics and self-reported vaccination status. These data were used to classify cases in different categories according to their vaccination status and whether they had a previous infection, as previously described*[3]*.

### BA.2 and BA.4/5 variant detection using S gene target failure

The TaqPath COVID-19 RT-PCR tests for three targets (S, ORF1ab and N) and is used by several laboratories in the Netherlands for SARS-CoV-2 diagnostics. S-gene target failure (SGTF) in combination with a proper signal (quantification cycle <= 30) from ORF1ab and N, also referred to as S gene not detected or S-dropout, has proven to be a highly specific proxy for SARS-CoV-2 variants containing the 69/70 deletion. Alpha[10, 11], Omicron BA.1[4], BA.4, and BA.5 variants possess the S 69/70 deletion[12], while the ancestral strains, Beta, Gamma, Delta[4], and Omicron BA.2[3] variants do not possess the deletion[12]. BA.4 and BA.5 cannot be distinguished using the TaqPath PCR, therefore we refer to SGTF as BA.4/5 and non-SGTF as BA.2. SGTF can only be used as a specific proxy for the variant when different variants containing or lacking the 69/70 deletion are not co-circulating or are only observed together for a very short period of time (see below). Whole genome sequencing (WGS) of a random selection of SGTF samples included in this study confirmed 52 (13.5%) BA.4 and 322 (83.6%) BA.5 cases among 385 sequenced SGTF cases. The proportion of BA.5 among SGTF cases increased over time. Non-SGTF was strongly associated with BA.2 (including BA.2 sub-variants; 485 of 495 (98.0%) cases during the study period).

### Variant detection of previous infections

Among the cases with a previous infection diagnosed in the community testing facility, the variant of the earlier infection was determined if a WGS or SGTF result was available (1609 out of 7625 reinfections (21.1%)). WGS was used to determine the previous variant in 117 of 1609 (7.3%) reinfections with variant data. For the other 1492 out of 1609 (92.7%), the variant was defined by SGTF result in combination with the testing date. Periods of successive >= 90% variant-specific S-detection confirmed by WGS were identified for the pre-VOC (18-01-2021 (start data collection) – 17-02-2021, non-SGTF), Alpha (18-01-2021 – 27-09-2021, SGTF), Delta (20-06-2021 – 07-01-2022, non-SGTF), Omicron BA.1 (23-11-2021 – 09-04-2022, SGTF), and Omicron BA.2 (29-01-2022 – end study, non-SGTF) variants (Fig S2). No periods were defined for the Beta and Gamma variants as they were indistinguishable from non-VOC, Delta and each other using SGTF with the >90% threshold.

### Statistical analysis

We compared the distribution of the following immune status groups between BA.2 and BA.4/5 infections: unvaccinated cases with and without a registered previous infection, primary vaccinated cases with and without a registered previous infection, booster vaccinated cases with and without a registered previous infection. We performed multinomial logistic regression to estimate the association between immune status, with unvaccinated without a previous infection as reference, and variants BA.2 and BA.4/5 based on SGTF, adjusting for testing week, age group (18-29, 30-49, 50-69, and 70+ years) and sex. If the Omicron BA.4/5 variants and BA.2 variants had similar ability to escape immunity from vaccination and/or previous infection, we would expect them to occur equally frequently in vaccinated and/or previously infected persons as in individuals without vaccination and previous infection, i.e. an odds ratio (OR) of 1. In addition, we performed multinomial logistic regression to estimate the association between previous infection (irrespective of previous variant) and previous variant status, with cases without a registered previous infection as reference, and variants BA.2 and BA.4/5 based on SGTF, adjusting for testing week, age group (18-29, 30-49, 50-69, and 70+ years) and sex.

Among cases with a previous infection, we calculated the interval between sample dates in days and tested difference in intervals between BA.2 and BA.4/5 infection with a Mann-Whitney test. The difference in distribution of previous variants found in reinfections between BA.2 and BA.4/5 was tested with a chi-square test with Monte Carlo simulated p-value and data was fit into 100 fields using the largest remainder method (for Fig 2B and Fig S3).

## Results

### Study population

Between 2 May and 24 July 2022, 19836 (73.8%) BA.4/5 and 7052 (26.2%) BA.2 cases were detected (Table 1). In the first week, BA.4/5 comprised 18/545 (3.3%) of cases and in the last week 2543/2572 (98.9%) (Fig. S1). BA.4/5 cases were generally younger than BA.2 cases (Table 1). Interestingly, the proportion of cases with a previous infection was larger among BA.4/5 cases overall (Table 1) and per week during this period (Fig. 1A). Of the 7052 BA.2 cases, 1408 (20.0%) were reinfections compared to 6217 out of 19836 (31.3%) BA.4/5 cases (Fig. 1A). No such differences were observed for vaccination statuses (Fig 1B).

**Fig. 1.**
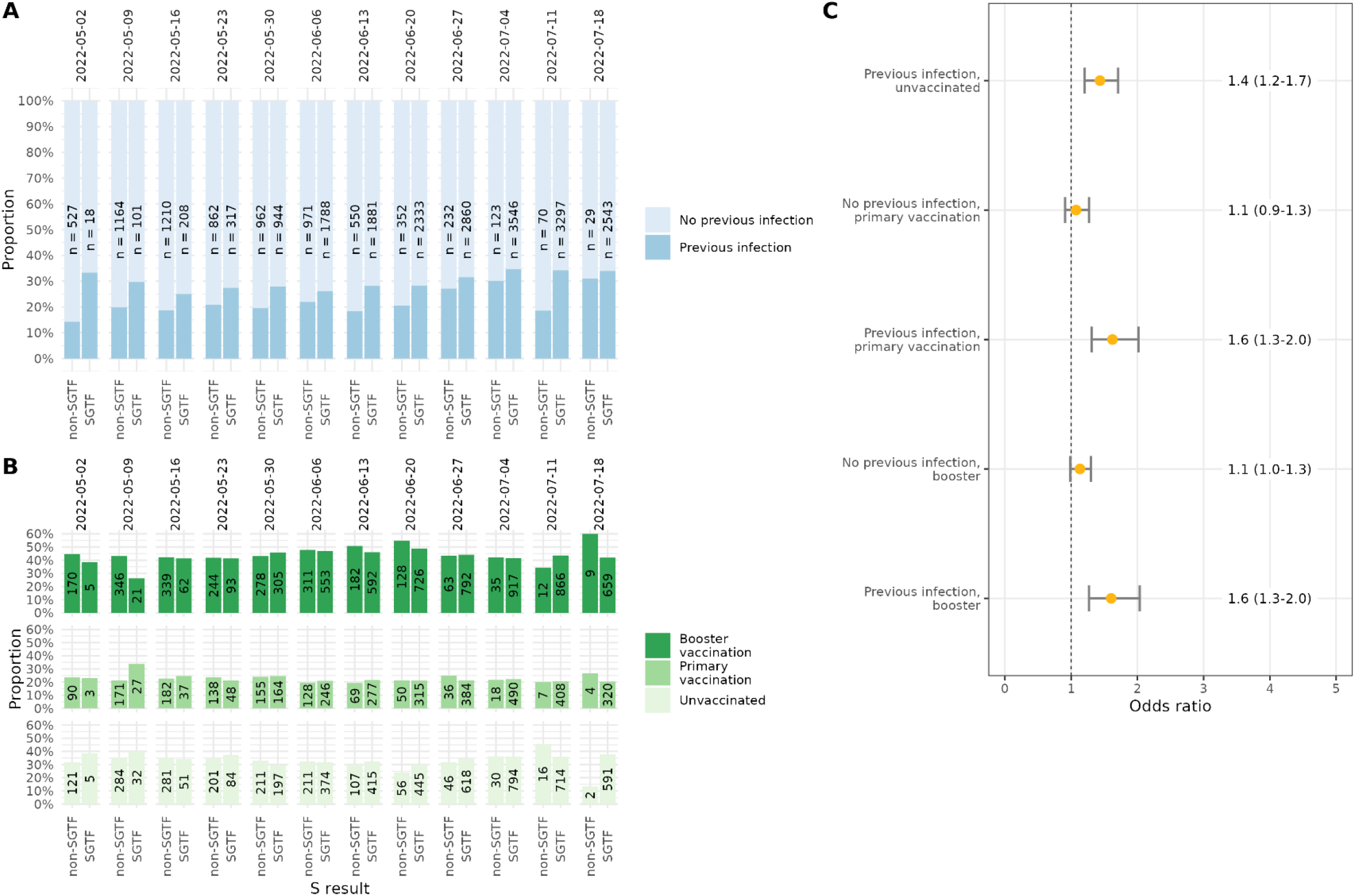
(A) Proportion of previous infections among non-SGTF (BA.2) and SGTF (BA.4/5) cases, n=26,888, the Netherlands, per week, 2 May - 24 July 2022. (B) Proportion of unvaccinated, primary, and booster vaccination among non-SGTF (BA.2) and SGTF (BA.4/5) cases, n=17,391. The number of cases corresponding to each group is displayed within the bars. (C) Association between previous infection and vaccination status and SGTF, with unvaccinated without a registered previous infection as reference group, n= 16,178, in individuals aged 18 years and older, adjusted for week of test, sex and age group (18-29, 30-49, 50-69, and 70+ years).

**Table 1.**
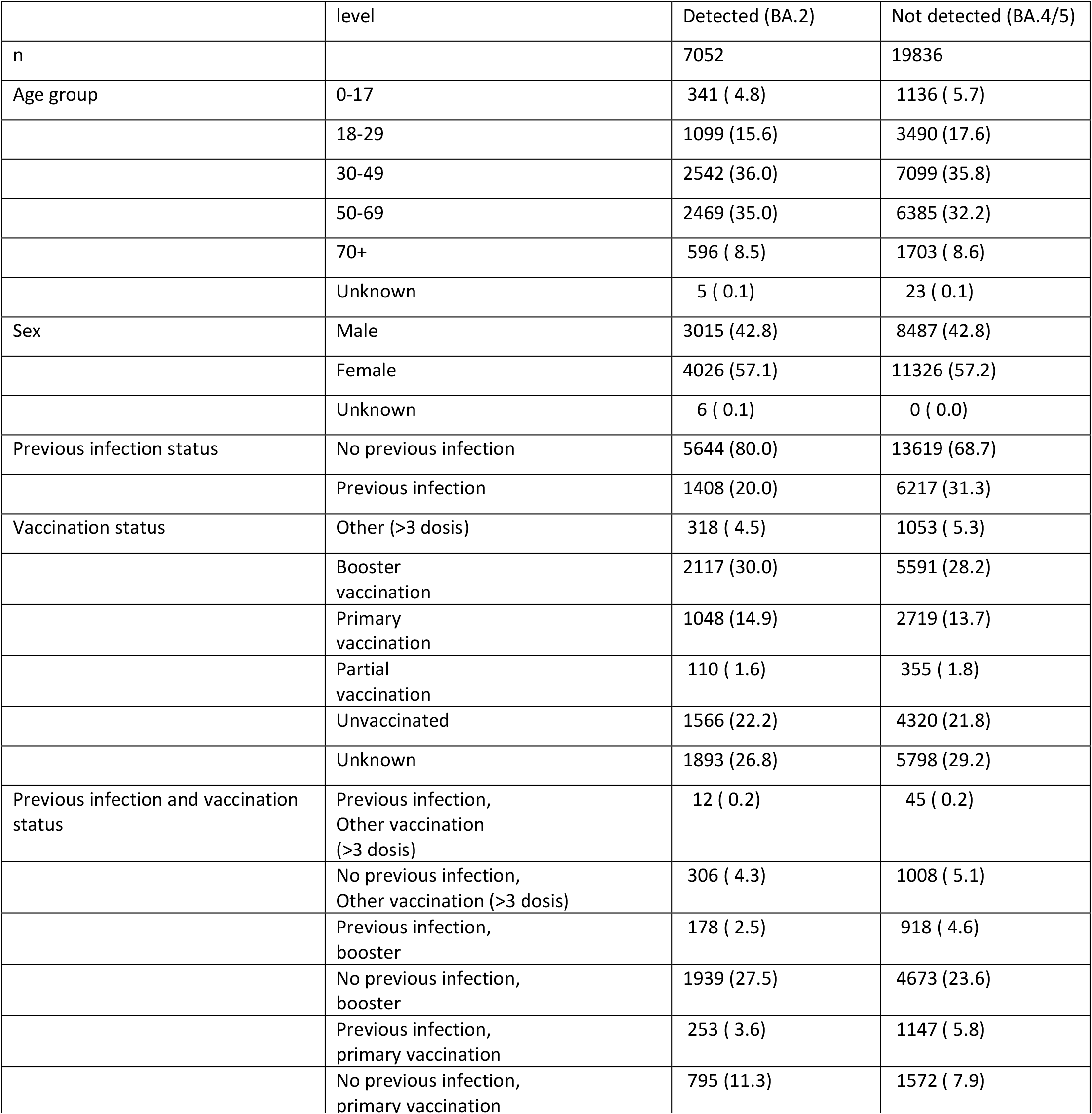
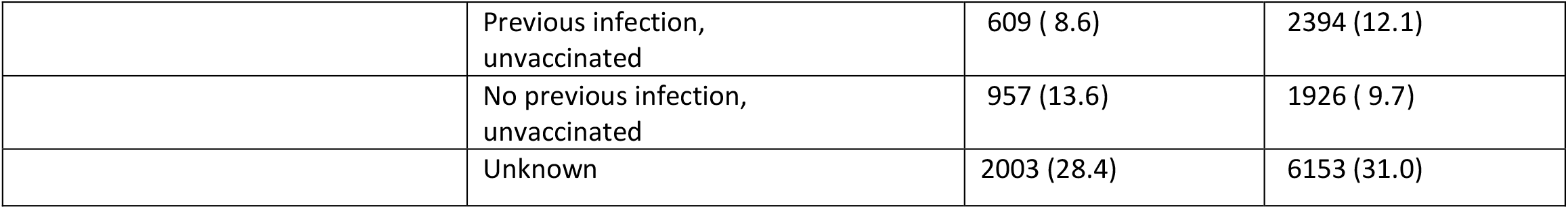
Characteristics of SARS-CoV-2 cases by non-SGTF (BA.2) and SGTF (BA.4/BA.5) status, n= 26,888, the Netherlands, 2 May - 24 July 2022

|  | level | Detected (BA.2) | Not detected (BA.4/5) |
| --- | --- | --- | --- |
| n |  | 7052 | 19836 |
| Age group | 0-17 | 341 ( 4.8) | 1136 ( 5.7) |
|  | 18-29 | 1099 (15.6) | 3490 (17.6) |
|  | 30-49 | 2542 (36.0) | 7099 (35.8) |
|  | 50-69 | 2469 (35.0) | 6385 (32.2) |
|  | 70+ | 596 ( 8.5) | 1703 ( 8.6) |
|  | Unknown | 5 ( 0.1) | 23 ( 0.1) |
| Sex | Male | 3015 (42.8) | 8487 (42.8) |
|  | Female | 4026 (57.1) | 11326 (57.2) |
|  | Unknown | 6 ( 0.1) | 0 ( 0.0) |
| Previous infection status | No previous infection | 5644 (80.0) | 13619 (68.7) |
|  | Previous infection | 1408 (20.0) | 6217 (31.3) |
| Vaccination status | Other (>3 dosis) | 318 ( 4.5) | 1053 ( 5.3) |
|  | Booster vaccination | 2117 (30.0) | 5591 (28.2) |
|  | Primary vaccination | 1048 (14.9) | 2719 (13.7) |
|  | Partial vaccination | 110 ( 1.6) | 355 ( 1.8) |
|  | Unvaccinated | 1566 (22.2) | 4320 (21.8) |
|  | Unknown | 1893 (26.8) | 5798 (29.2) |
| Previous infection and vaccination status | Previous infection, Other vaccination (>3 dosis) | 12 ( 0.2) | 45 ( 0.2) |
|  | No previous infection, Other vaccination (>3 dosis) | 306 ( 4.3) | 1008 ( 5.1) |
|  | Previous infection, booster | 178 ( 2.5) | 918 ( 4.6) |
|  | No previous infection, booster | 1939 (27.5) | 4673 (23.6) |
|  | Previous infection, primary vaccination | 253 ( 3.6) | 1147 ( 5.8) |
|  | No previous infection, primary vaccination | 795 (11.3) | 1572 ( 7.9) |
|  | Previous infection,<br>unvaccinated | 609 ( 8.6) | 2394 (12.1) |
|  | No previous infection,<br>unvaccinated | 957 (13.6) | 1926 ( 9.7) |
|  | Unknown | 2003 (28.4) | 6153 (31.0) |

### Immune status and BA.4/5 vs. BA.2

The higher proportion of reinfections among BA.4/5 cases remained after adjustment for age, sex and calendar time and the effect was present in unvaccinated and vaccinated cases (OR ranging from 1.4 (95% CI: 1.2-1.7) in unvaccinated cases to 1.6 (95% CI: 1.3-2.0) in cases with primary or booster vaccination; Fig. 1C). Among cases without a previous infection, there was no association between vaccination status and BA.4/5 (OR of 1.1 for primary and booster vaccination; Fig. 1C).

### Reinfections

Overall, previous infection was associated with an increased risk of BA.4/5 infection compared with the risk for infection with BA.2, after adjustment for age, sex and calendar time with an OR of 1.4 (95% CI: 1.3-1.5; Fig 2A top panel). Among reinfected cases, intervals between infections were shorter in BA.4/5 cases (median interval 182 days) compared to BA.2 cases (median interval 206 days, *p-*value 0.004, Fig 2C). A likely explanation for this observation is that BA.4/5 cases more frequently had a previous BA.1 infection than BA.2 cases (63.1% and 46.4%, respectively, p<0.001, Fig 2B). The large number of previous BA.1 infections can partially be explained by the large pool of individuals with BA.1-induced immunity. Nonetheless, this would not result in a difference in previous BA.1 infections among current BA.2 and BA.4/5 cases. In addition, also stratified by calendar time of the current infection, the proportion of previous BA.1 infections is higher among BA.4/5 cases than BA.2 cases (Fig S3), indicating higher levels of immune escape by BA.4/5 from protection conferred by BA.1 infection compared to other previous infections. Indeed, when adjusting for age, sex and calendar time, particularly previous BA.1 infection was associated with BA.4/5 (OR: 1.9 (95% CI: 1.5-2.6). For all other previous variants, except BA.2, a trend towards an increased risk of BA.4/5 compared to BA.2 was observed (Fig. 2A bottom panel).

**Fig. 2.**
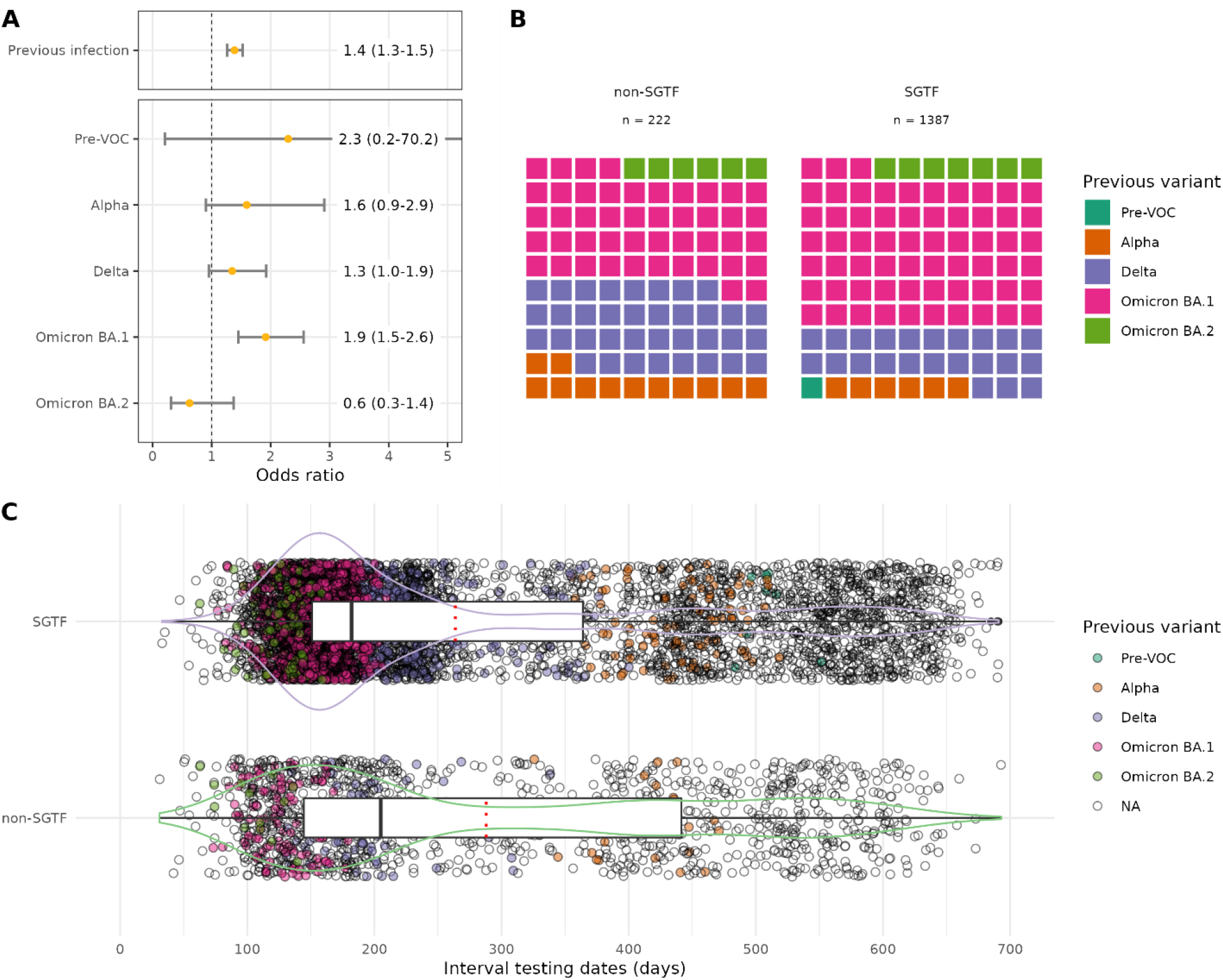
(A) Association between previous infection (n= 25,377, top panel), or previous variant (n = 19,895, bottom panel), and SGTF, with no registered previous infection as reference group. Analyses are in individuals aged 18 years and older, adjusted for week of testing, sex and age group (18-29, 30-49, 50-69, and 70+ years). (B) Distribution of previous variants among reinfection pairs with a known previous variant for SGTF (BA.4/5) and non-SGTF (BA.2) cases. Figure stratified by time in Fig S3. (C) Interval of testing dates in reinfections of non-SGTF (BA.2) and SGTF (BA.4/5) cases in days (n = 7625). Reinfections are colored to indicate the variant of the previous infection. Median is displayed in black (line) and mean in red (dotted).

## Discussion

Evidence on the presence of escape of infection- and/or vaccination-induced immunity by novel SARS-CoV-2 VOCs is highly relevant for vaccine policy. We found that Omicron BA.4/5 cases more often had previous infections than BA.2 cases, even when adjusted for week of testing and irrespective of vaccination status. This indicates relatively more evasion of infection-induced immunity by BA.4/5. There was no association between vaccination status and BA.4/5 infection versus BA.2 infection, suggesting that there is equal protection induced by vaccination against the BA.4/5 and BA.2 variants. Among reinfection cases, we found shorter intervals for BA.4/5 than for BA.2 infections, whereby the previous infection was more often BA.1. This suggests that BA.1-induced immunity protects less well and/or shorter against BA.4/5 infection than against BA.2 infection.

*In vitro* studies have shown BA.4/5 can escape neutralizing antibodies elicited by vaccination and Omicron (BA.1 or BA.2) infection[7-9]. A pre-print from Denmark showed no difference in vaccine effectiveness between BA.5 and BA.2[13], which is in line with our results. This study observed a protective effect of previous Omicron BA.1/2 infection of 93.6% (95% CI: 92.1-94.8) and 96.3% (95% CI: 95.8-96.7) against BA.5 and BA.2 infection, respectively. The difference between these effects, a ratio of 1.7, corresponds with our results, as we found an OR of 1.9 for Omicron BA.1 infection. Also for a previous Alpha infection our results are similar, where the Danish study observed protection estimates of 65.4% (95% CI: 49.8-76.2) and 74.5% (95% CI: 68.7-79.2) for BA.5 and BA.2, a ratio of 1.4. However, for previous Delta infections our estimates differ, as the Danish study observed protection estimates of 46.9% (95% CI: 27.0-61.3) and 77.2% (95% CI: 72.2-81.3) for BA.5 and BA.2, a ratio of 2.3, while we found an OR of 1.3 [13]. A pre-print from Portugal reported similar results regarding the difference between BA.4/5 and BA.2 in protection from a previous infection without vaccination (OR: 1.8, 95% CI: 1.3-2.5) or with primary vaccination (OR: 1.7, 95% CI: 1.4-2.0) both relative to unvaccinated without a previous infection[14]. However, different findings were made for booster vaccination with a previous infection, as they did not find a statistically significant reduction (OR: 1.2, 95% CI: 1.0-1.5). Similar to our results, vaccination status irrespective of previous infection status did not differ between BA.4/5 and BA.2 cases in the UK and Portugal[14, 15].

Overall, a reduction in protection conferred by previous infection is observed for BA.4/5 relative to BA.2 and no changes in protection by vaccination are observed. The reduction in protection from previous infection seems mainly driven by escape from BA.1-induced protection. Others argued that the large number of reinfections with BA.5 is caused by the large pool of BA.1 infections[16]. However, we show that also immune escape plays a role in the large number of reinfections with BA.5. Still, the difference in escape from previous infection for BA.2 and BA.4/5 are smaller in comparison to differences found for BA.1 and Delta[4], suggesting more escape between VOCs than within the Omicron lineage.

A case-only analysis indicates whether protection against a variant changes relative to the reference variant[17]. An advantage of our case-only approach on comparing levels of protection between variants is that it prevents bias from poor control selection or bias from exposure misclassification differential by disease status. Previous studies used a similar design and found relative differences between earlier VOCs[4, 18, 19]. However our study has some limitations. First, a considerable but unknown part of the previous infections will not have been registered because of lack of testing or symptoms, and this will have misclassified some individuals as not previously infected. This will likely have diluted the actual differences in protection between previously and not previously infected individuals. Second, self-reported vaccination status could have led to some misclassification, however this is not likely to differ between variants. Third, we could not distinguish BA.4 from BA.5 using TaqPath-PCR data, although the majority of the SGTF cases were BA.5 infections (83.6%) and the Spike sequences of BA.4 and BA.5 are identical, therefore no differences in anti-S antibody escape are expected[9]. However, our results might differ slightly from estimates specific for BA.5 as 16.4% of the variants found among SGTF are BA.4 and other variants. Limiting our analysis to WGS data only would give us insufficient power due to small sample size. In addition, previous variants classified with TaqPath could sometimes be misclassified as we used a 90% threshold. This is unlikely to meaningfully affect our results.

In general, protection conferred by previous infection is good, especially in combination with vaccination[3, 13, 16]. Our results suggest a reduction in protection from previous infection against BA.4/5 compared with the BA.2 variant. This immune evasion is also observed within the Omicron lineage and specifically for the first Omicron lineage that became dominant, BA.1. This is of importance, considering the relatively large fraction of the population having BA.1-induced immunity in comparison to other variants. In contrast, no association between vaccination status and BA.4/5 infection versus BA.2 infection was observed, indicating a similar vaccine effectiveness against infection between BA.2 and BA.4/5. Immune evasion within the Omicron lineage allows for repeat Omicron infections to occur, and is thereby informative for considerations on vaccine updates and stresses the importance of studies on immune evasion by current and novel variants.

## Data Availability

Data produced in the present study are available upon reasonable request to the authors. Restrictions apply to the availability of case-based data.

## Acknowledgements

The authors would like to thank all personnel at the 25 Public Health Services for data collection in the national surveillance database, the members of the RIVM genomic surveillance team, and the members of the RIVM COVID-19 surveillance and epidemiology team.

## Ethical statement

The Centre for Clinical Expertise at the National Institute for Public Health and the Environment (RIVM) assessed the research proposal following the specific conditions as stated in the law for medical research involving human subjects. The work described was exempted for further approval by the ethical research committee. Pathogen surveillance is a legal task of the RIVM and is carried out under the responsibility of the Dutch Minister of Health, Welfare and Sports. The Public Health Act (Wet Publieke Gezondheid) provides that RIVM may receive pseudonymised data for this task without informed consent.

## Contributions

HV and IvW analyzed the sequence data. NvM, NEK and SPA performed data analysis. SPA performed statistical analysis, and MJK supervised statistical analysis. BdG, HEM, SJMH, SvdH, DE, MJK contributed analytically to the interpretation of the data. SPA drafted the manuscript under supervision of DE and MJK with input from all authors. All authors reviewed the manuscript and approved the final version.

