## Supplemental information for "Higher risk of SARS-CoV-2 Omicron BA.4/5 infection than of BA.2 infection after previous BA.1 infection, the Netherlands, 2 May to 24 July 2022"

### Supplement figures and Tables:

Fig S1

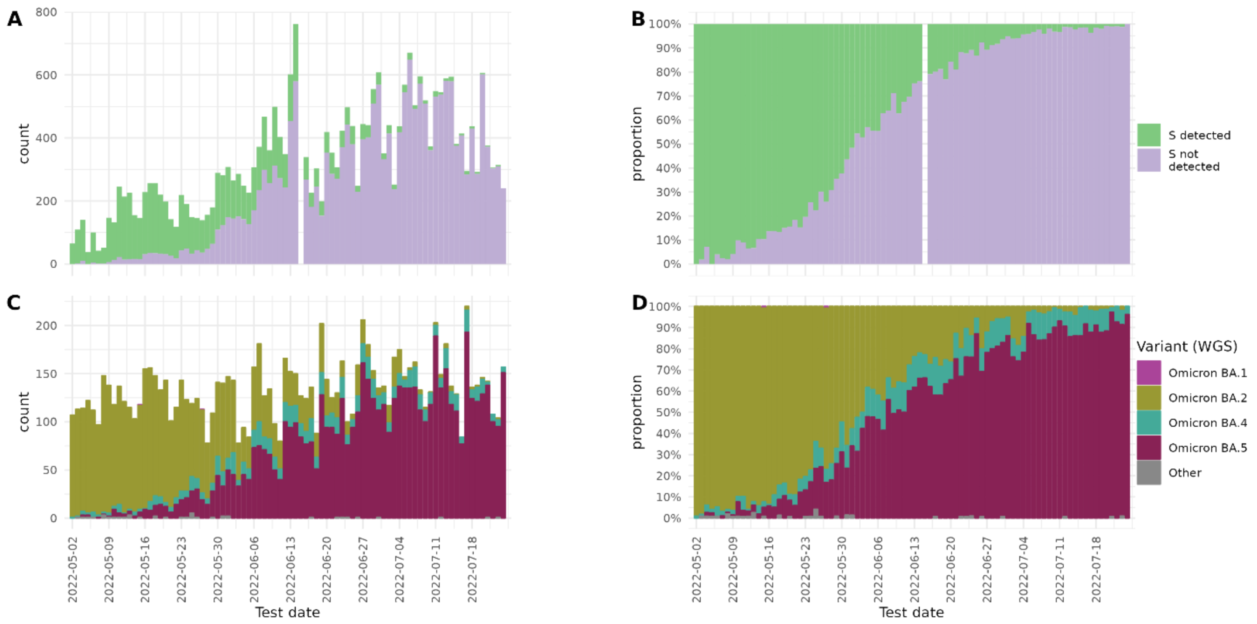


Fig S1: SGTF status and WGS typed variant found in community surveillance, the Netherlands, 2 May - 24 July 2022. Number (A) and proportion (B) of S-gene target failure (SGTF) and non-SGTF positive tests over time (n = 26888). Number (C) and proportion (D) of whole genome sequencing (WGS) typed variant found in national genome surveillance of community testing samples in the Netherlands (n = 11249).

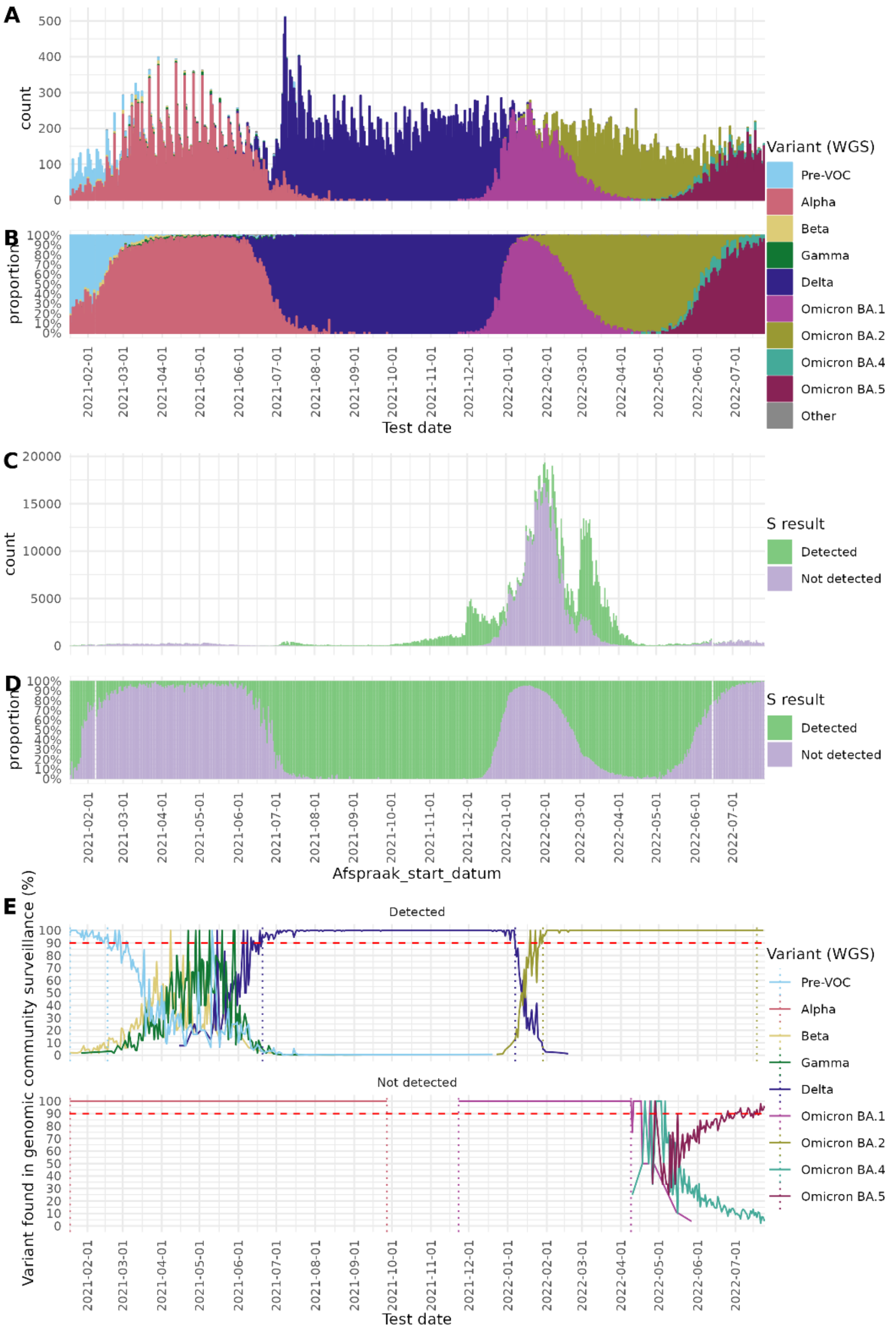


Fig S2 detection method of variant using SGTF. (A) number and (B) proportion of WGS variants found per date of testing. (C) number and (D) proportion of SGTF result detected per date of testing. (E) Proportion of the whole genome sequencing (WGS) typed variants found in national genome surveillance of community surveillance by SGTF status of the variant. Horizontal red striped line indicates the 90% threshold. Vertical dotted lines indicate the time periods used for previous variant detection using SGTF.

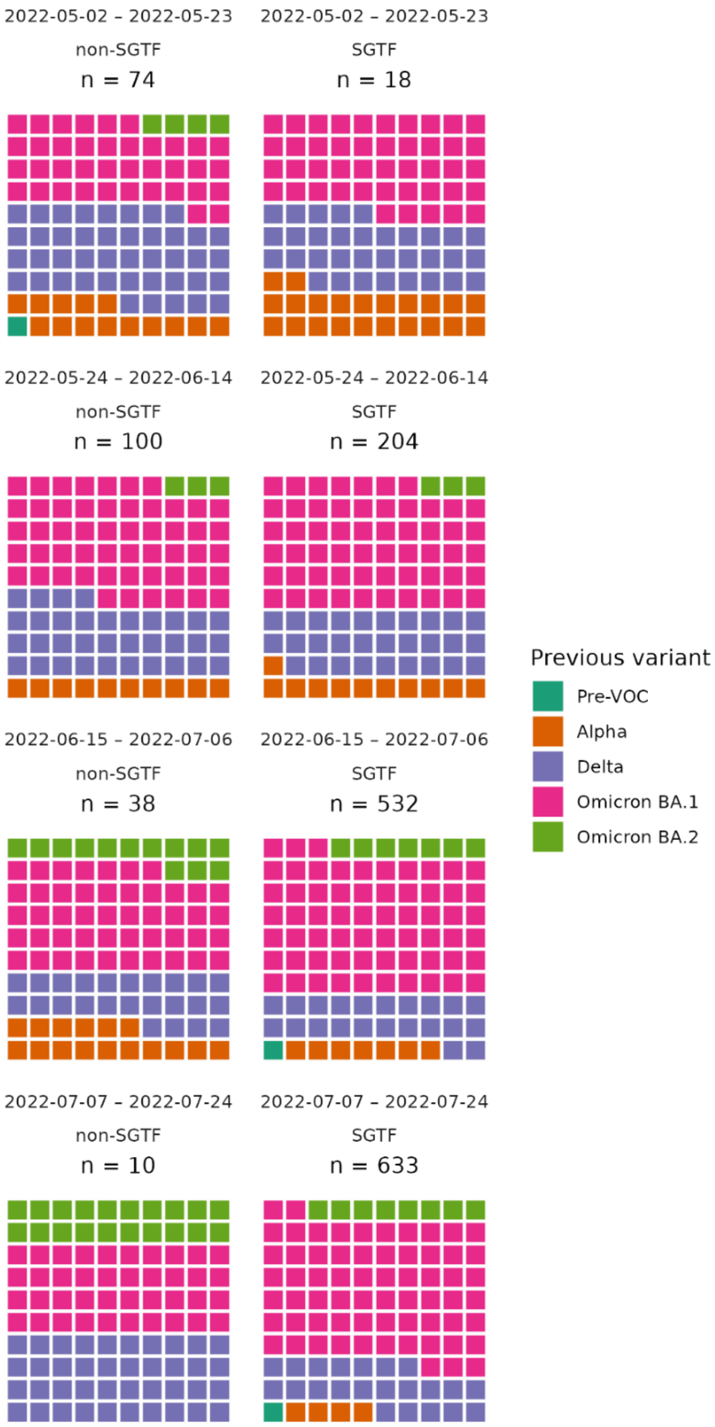


Fig S3 *Proportion of previous variants among reinfection pairs with a known previous variant for SGTF (BA4.5) and non-SGTF (BA.2) cases stratified by testing date (3 week groups).*
